# Integration of single-cell and bulk RNA sequencing reveals TREM1 as a promising biomarker and therapeutic target for gouty arthritis

**DOI:** 10.64898/2026.05.15.26353351

**Authors:** Wei Jinfeng, Zheng Jiarui, Qiu Hongbin

## Abstract

**Objective:** This study aimed to systematically screen for potential candidate biomarkers and identify therapeutic targets associated with gouty arthritis (GA) through integrated analyses of single-cell and bulk RNA sequencing (RNA-seq) data.

**Methods:** The single-cell dataset GSE211783 and the bulk RNA-seq dataset GSE160170 were analyzed using a series of bioinformatic approaches, including cell clustering, differential expression analysis, immune cell infiltration assessment, protein-protein interaction network construction, gene set enrichment analysis, as well as drug sensitivity evaluation. To establish an animal model of GA, monosodium urate crystals were injected intra-articularly into experimental mice. Joint swelling was evaluated, and morphological changes in joint tissues were analyzed through hematoxylin–eosin staining. The presence of TREM1-positive cells was detected by immunohistochemistry and the level of TREM1 protein expression in joint tissues were assessed by Western blotting.

**Results:** We identified 102 differentially expressed genes (DEGs) and 14 signaling pathways associated with GA. The PPI network revealed 25 hub genes, of which 17 (including TREM1, TNF, PTGS2, and NLRP3) were highly expressed and 8 (including FCGR3B and CXCR6) showed low expression in the GA samples. These genes correlated significantly with the infiltration levels of macrophages. Among the hub genes, TREM1 was selected for further validation because it correlated significantly with all 14 differential pathways. In animal experiments, GA mice developed marked joint swelling and inflammatory tissue injury, along with a significant increase in TREM1-positive cells and TREM1 protein expression.

**Conclusion:** Integrative analysis of single-cell and bulk RNA-seq data identified 102 GA-related DEGs and 14 key pathways, from which 25 hub genes were screened. TREM1 is significantly upregulated in GA and may be linked to macrophage function, providing new insights into biomarker and therapeutic target discovery for GA.

## 1. Introduction

Gouty arthritis (GA) represents a sterile autoimmune form of arthritis driven by monosodium urate (MSU) crystal accumulation in joints and surrounding connective tissues. It arises from disordered purine metabolism, resulting in either decreased excretion or excessive production of uric acid, which in turn elevates serum uric acid levels ^[1, 2]^ . Patients with GA commonly experience sudden and severe joint pain, together with swelling and redness of the affected joints. The rising incidence of this condition, most prominently in developed countries, along with its frequent coexistence with cardiovascular disease, metabolic syndrome, and chronic kidney disease, has rendered it a notable public health concern ^[3, 4]^. Given the complex pathophysiology of GA, numerous drugs have been developed for its clinical management. Non-steroidal anti-inflammatory drugs, glucocorticoids, and oral colchicine currently represent the first-line therapeutic options for GA ^[5]^ . However, these conventional therapies do not consistently achieve satisfactory clinical outcomes. Furthermore, they may induce significant adverse effects, including severe hepatic and renal toxicity, gastrointestinal reactions, and other complications, which restrict their clinical use ^[6]^ . With the advancement of precision medicine and a deeper understanding of disease heterogeneity, various biologics have gradually emerged ^[7, 8]^. However, there remains a lack of sufficient data regarding the long-term safety of these biologics, the optimal timing for discontinuation, and comparative studies between different biologics. Therefore, further identification of biomarkers and therapeutic targets will facilitate the optimization of clinical treatment strategies for GA, as well as improve treatment efficacy and patient prognosis.

Currently, GA is widely recognized as a classic inflammatory disease triggered by stimulation of the innate immune system. It is characterized by the participation of multiple types of immune cells, including lymphocytes, monocytes/macrophages, and neutrophils, whose phenotypic conversion is crucial in the onset and progression of GA ^[9, 10]^. Macrophages are among the earliest recognized and most extensively studied immune cells, playing a key role in regulating the entire course of GA ^[11, 12]^. Numerous studies have confirmed that MSU crystals promote the phenotypic shift of macrophages towards the M1 phenotype ^[13, 14]^. M1 macrophages secrete inflammatory cytokines such as interleukin (IL)-1βand IL-18, which further enhances inflammatory responses and mediate the development of intra-articular inflammation ^[15]^. During the resolution phase of inflammation, M2 macrophages progressively predominate and secrete factors that suppress inflammatory responses as well as inflammation ^[16]^. The M1-to-M2 macrophage transition2 may represent a self-limiting step in the malignant progression of acute inflammation. Accordingly, reducing M1 macrophages and the production of their associated cytokines may help alleviate inflammation in GA and its related complications ^[17]^.Therefore, a better understanding of macrophage-associated marker genes in GA is beneficial for modulating macrophage phenotypes, controlling disease progression, and contributing to the identification of new therapeutic strategies for GA. Consequently, this study adopts a macrophage-centric approach, integrating single-cell and bulk RNA sequencing (RNA-seq) data to screen for candidate biomarkers as well as potential targets for therapeutic intervention in GA.

## 2. Materials and Methods

### 2.1 Data retrieval and pre-processing

The datasets GSE211783 (a single-cell transcriptomic dataset) and GSE160170 (a bulk RNA-Seq dataset) were downloaded from the Gene Expression Omnibus (GEO) database (https://www.ncbi.nlm.nih.gov/geo/). Gene names for the probes on the GSE160170 chip were annotated by applying the idmap3 package in R (version 4.2.2).

### 2.2 Analysis of single-cell data

The ‘Read10X’ function in the Seurat package in R (version 4.2.2) was applied to read single-cell data, and low-quality single-cell sequencing results were filtered out based on ‘n_feature < 5000’ (the number of genes in a single cell) and ‘percent.mt < 10’ (the proportion of mitochondrial genes in a single cell). The integration methods provided by the ‘FindIntegrationAnchors’ function in the Seurat package in R (version 4.2.2) was employed for the removal of batch effects. These methods first facilitated the identification of pairs of cells across datasets in matching biological states (anchors), and then the correction for batch effects between samples based on these anchors. Seurat constructed a K-nearest neighbour graph to connect cells with similar gene expression patterns through a graph-based clustering approach, and then attempted to partition this graph into highly interconnected ‘clusters’. The FindClusters function was used to perform graph-based clustering, with the resolution parameter set to 0.3. Concurrently, the FindMarkers function was utilized to screen for differentially expressed genes (DEGs) within the cell clusters. Known cell marker genes retrieved from the cellMarker2 database (http://117.50.127.228/CellMarker/) were used for the annotation of the 21 identified cell clusters.

### 2.3 Transcriptomic data analysis

The limma package implemented in R (version 4.2.2) was employed for performing differential analysis of the GSE160170 dataset (encompassing 6 healthy controls and 6 gout patients). DEGs were identified based on P-values < 0.05 and |logFC| < 1. The CIBERSORT package implemented in R (version 4.2.2) was applied for conducting immune cell infiltration analysis, and a t-test (p < 0.05) was applied to select genes showing differential immune cell infiltration.

### 2.4 Screening for intersecting genes

An intersection analysis was performed between macrophage marker genes from the single-cell transcriptomic dataset GSE211783 and DEGs from the bulk RNA-seq dataset GSE160170, and the results were visualized.

### 2.5 Gene set variation analysis (GSVA) pathway enrichment scores

The Kyoto Encyclopedia of Genes and Genomes pathway background gene set c2.cp.kegg_legacy.v2023.2.Hs.symbols.gmt was downloaded from MSigDB (https://www.gsea-msigdb.org/gsea/msigdb/). Pathway activity scores were predicted using the gsva package in R (version 4.2.2) based on the GSE160170 dataset. Subsequently, the limma package in R (version 4.2.2) was applied to screen for pathways with significantly different scores between disease and control samples (p < 0.05 and |logFC| > 2).

### 2.6 Construction of a PPI network and identification of core genes

STRING (https://cn.string-db.org/) was employed for predicting interactions among the identified intersecting genes, resulting in the selection of 815 gene pairs with a composite score greater than 0.4. The igraph package in R (version 4.2.2) was applied for constructing the PPI network. The top 20 core genes were calculated using the MCC, MNC, and DMNC algorithms in the cytoHubba plugin of Cytoscape, and the genes selected by all three algorithms were combined to form the hub genes.

### 2.7 Drug sensitivity prediction

oncoPredict is a drug prediction model developed based on the Gene Disruption Screen Collection (GDSC) database. GDSC comprises expression matrices for approximately 20,000 genes across more than 800 cell lines, as well as IC_50_ values for approximately 200 drugs across the same 800+ cell lines. We utilized the oncoPredict package in R (version 4.2.2) to analyze the correlation between hub genes and drug sensitivity.

### 2.8 Animal model generation

Male wild-type C57BL/6 mice (n = 16; 6–8 weeks old; 20 ± 0.5 g) were supplied by Liaoning Changsheng Biotechnology Corporation Limited. Prior to experimentation, the mice were acclimatized for seven days under specific pathogen-free conditions, maintained at 18–22 °C on a 12-hour light/12-hour dark cycle, and provided with unrestricted access to food and water throughout this period. This study’s animal experimental procedures complied with the Guidelines for the Care and Use of Laboratory Animals and received approval from the Biomedical and Medical Ethics Committee of the School of Public Health, Jiamusi University (Approval No. 20240318093512277577318). A solution was prepared by dissolving uric acid (1 g) and sodium hydroxide (0.5 g) in double-distilled water (ddH_2_O; 100 mL). The pH of the prepared solution was adjusted to 7.4 using hydrochloric acid to ensure complete saturation of sodium urate. The mixture was kept at 4°C overnight to facilitate crystallization. Subsequently, MSU crystals were obtained by centrifugation (3600 rpm, 5 min, 4 °C). The collected crystals were washed twice with ddH_2_O, dried, autoclaved (30 min, 130 °C), and weighed under sterile conditions. The crystals were ultimately suspended in phosphate-buffered saline (PBS) to obtain a 20 mg/mL suspension. Mice were randomly allocated to each of two groups (control and model; n=8 per group). Mice in the model group received an injection of MSU crystal suspension supplemented with 0.5 mg of MSU into the right hind paw joint, while those in the control group were injected with the same volume of sterile PBS at the same site. 3 h after MSU injection, a digital caliper was used to measure joint swelling, and the swelling index was subsequently calculated as follows: swelling index = (measured circumference - baseline circumference) / baseline circumference. 72 h after MSU injection, the mice were anaesthetized and euthanized by cervical dislocation; synovial fluid and joint tissue were then collected for subsequent experiments.

### 2.9 Hematoxylin-eosin (HE) staining

To assess the morphology of mouse joint tissues, HE staining was applied. Joint samples were fixed, decalcified, paraffin-embedded, and then sectioned into slices (4 μm-thick). Following dewaxing and rehydration, hematoxylin staining was performed for approximately 10 min, followed by 15 min of rising under running water to remove excess dye. Subsequently, the sections were stained with eosin for 2 min, and excess stain was washed off with tap water. After dehydration and clearing, the tissue sections were mounted with neutral resin, and the tissue morphology was observed under a microscope, with images recorded.

### 2.10 Immunohistochemistry (IHC)

IHC was used to detect TREM1 expression in mouse joint tissue. Mouse joint tissue sections were subjected to antigen retrieval in 0.01 M citrate buffer (pH 6.0). Endogenous enzymes were inactivated by treatment with 3%H_2_O_2_ for 10 min at room temperature. After washing with PBS, sections were subjected to an overnight incubation at 4 °C with appropriately diluted primary antibody. The following day, sections were incubated with secondary antibody (100 μL) for 30 min at 37 °C. Subsequently, visualization was performed using the DAB method and counterstaining was carried out using haematoxylin. After dehydration and mounting, positive cells in the tissue were carefully observed under a microscope.

### 2.11 Western blot

Mouse joint tissue samples were transferred to a pre-chilled tissue homogenizer and homogenized at 3,000 rpm for 60 s. RIPA lysis buffer containing protease inhibitors was added to the tissue homogenate, followed by thorough mixing and incubation on ice for 15 min. The mixture was centrifuged (10,000 rpm; 15 min; 4 °C), and the resulting supernatant was collected. Quantification of the total protein was carried out using a BCA assay kit. Following separation by SDS-PAGE, proteins were transferred to a polyvinylidene fluoride membrane (Millipore, USA). The membrane was next blocked for 1 h with TBST containing 5% skimmed milk powder at room temperature. Next, the membrane was incubated first with the primary antibody (overnight; 4 °C) and then with HRP-labelled secondary antibody (1 h). WB signals were visualized using an ECL kit, and analysis of protein bands were performed with ImageJ software.

### 2.12 Statistics and analysis

Bioinformatics analysis was performed using R (version 4.4.1). Statistical analyses of the experimental results were conducted using GraphPad Prism 8. All experimental data are reported as mean ± standard deviation 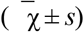. Group comparisons were analyzed with a two-tailed Student’s t-test, with statistical significance set at P < 0.05.

## 3. Results

### 3.1 Results of single-cell data analysis

The results of the single-cell dataset GSE211783 before and after quality control are shown in Figure 1A. After merging and batch-normalizing the data, the results are shown in Figure 1B. Following clustering of the cellular data, the dimension-reduced scatter plot is shown in Figure 1C, yielding a total of 21 cell clusters. After annotation using CellMarker2, eight cell types were identified: B_cell, endothelial cell, Macrophages, Monocyte, NK_cell, Platelets, Pro-Myelocyte, and T_cells (Figure 1D).

**Figure 1.**
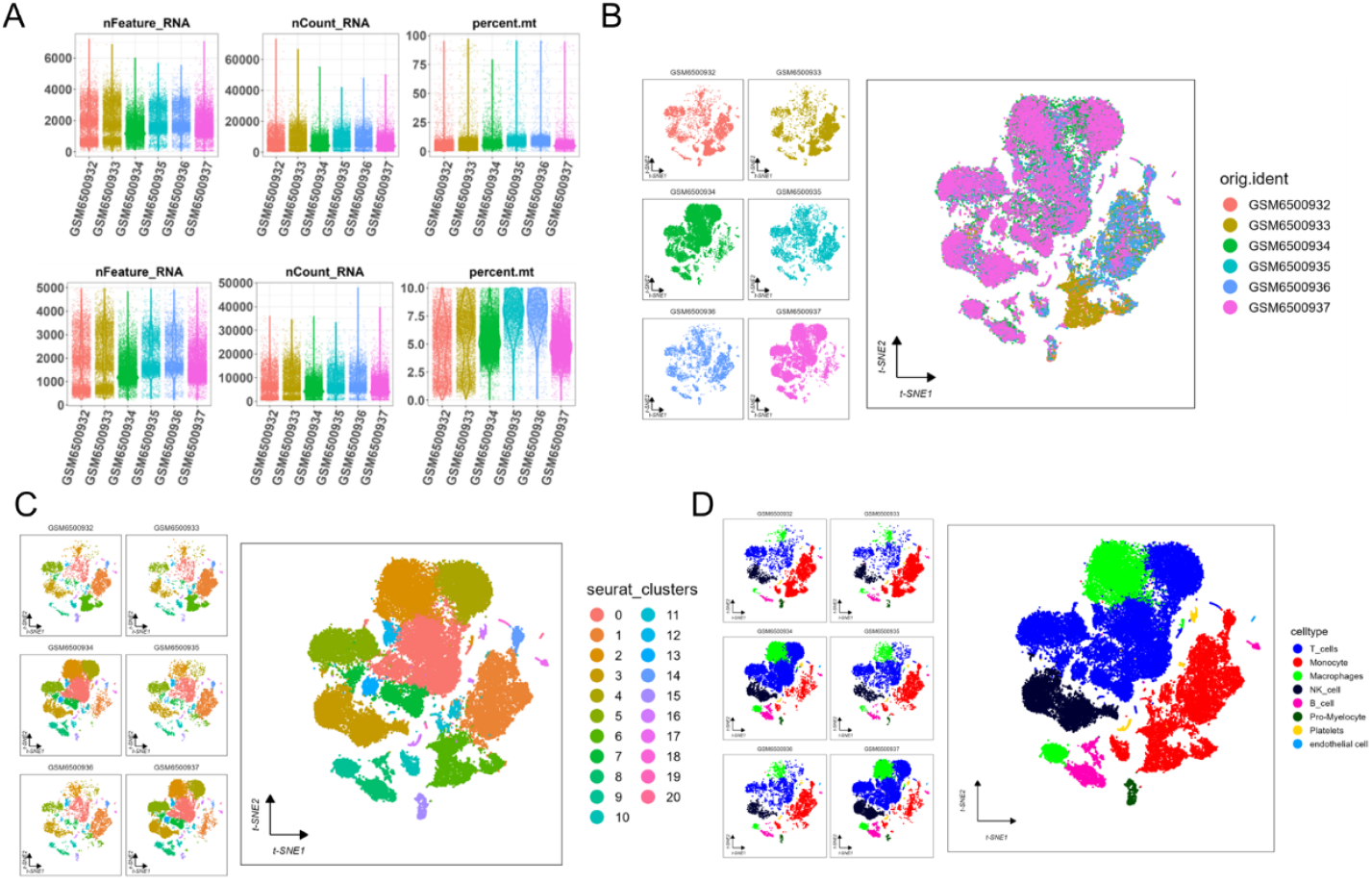
Analysis results of the single-cell sequencing dataset. A: Violin plots before (top) and after (bottom) data quality control; B: tSNE-dimension-reduced scatter plot after data merging and batch effect correction; C: tSNE-dimension-reduced scatter plot of cell clustering; D: tSNE-dimension-reduced scatter plot of cell annotation.

### 3.2 Analysis of marker gene expression in various cell types within the single-cell dataset

As illustrated in Figure 2A, the top 20 marker genes from each cell cluster exhibit distinct expression patterns across the eight cell types: B cells, monocytes, platelets, NK cells, macrophages, endothelial cells, T cells, and pro-myelocytes. Overall, the top 20 marker genes from each cell cluster exhibited relatively high expression in B cells and macrophages. Figure 2B presents the expression patterns of the top 20 macrophage marker genes across the eight cell types. These marker genes also exhibited higher overall expression in B cells and macrophages; however, the proportion of cells expressing these genes within the overall B cell and macrophage populations is limited, and not all B cells and macrophages express these genes.

**Figure 2.**
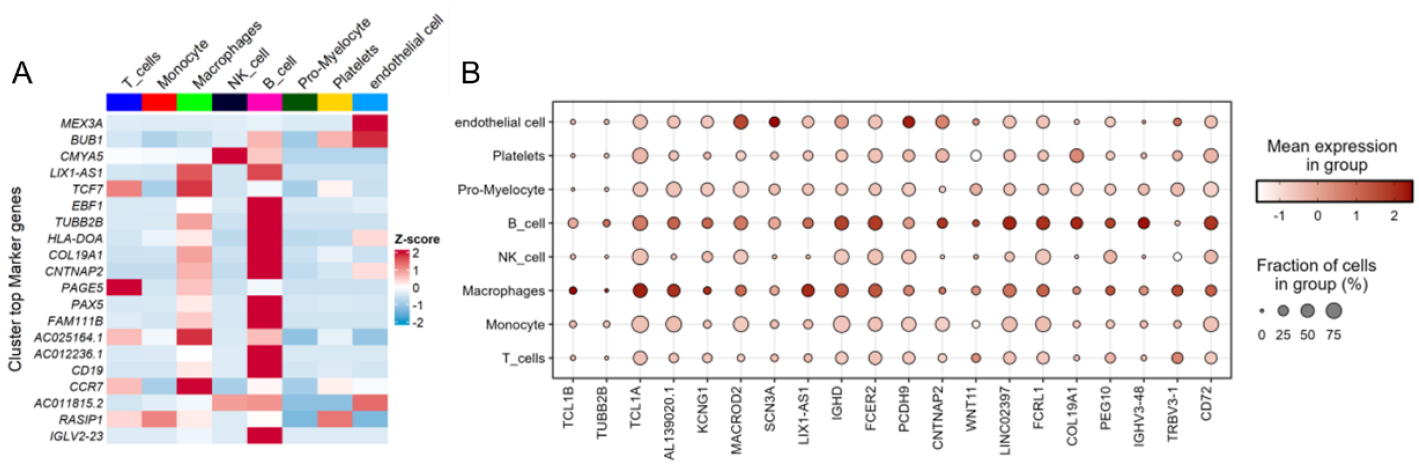
Analysis of marker gene expression across cell types in the single-cell dataset. A: Heatmap showing the top 20 marker genes in cell clusters across the eight cell types; B: Bubble plot illustrating the top 20 macrophage marker genes across the eight cell types.

### 3.3 Analysis results of the bulk RNA dataset

910 DEGs were screened out from the bulk RNA-seq dataset GSE160170 (Figures 3A–B). The immune infiltration patterns across different cell types in each sample are shown in Figure 3C. Significant differences in infiltration levels were observed for Naïve B cells, M1 macrophages, plasma cells, activated dendritic cells, and follicular helper T cells between the disease and control groups (*P* < 0.05, Figure 3D).

**Figure 3.**
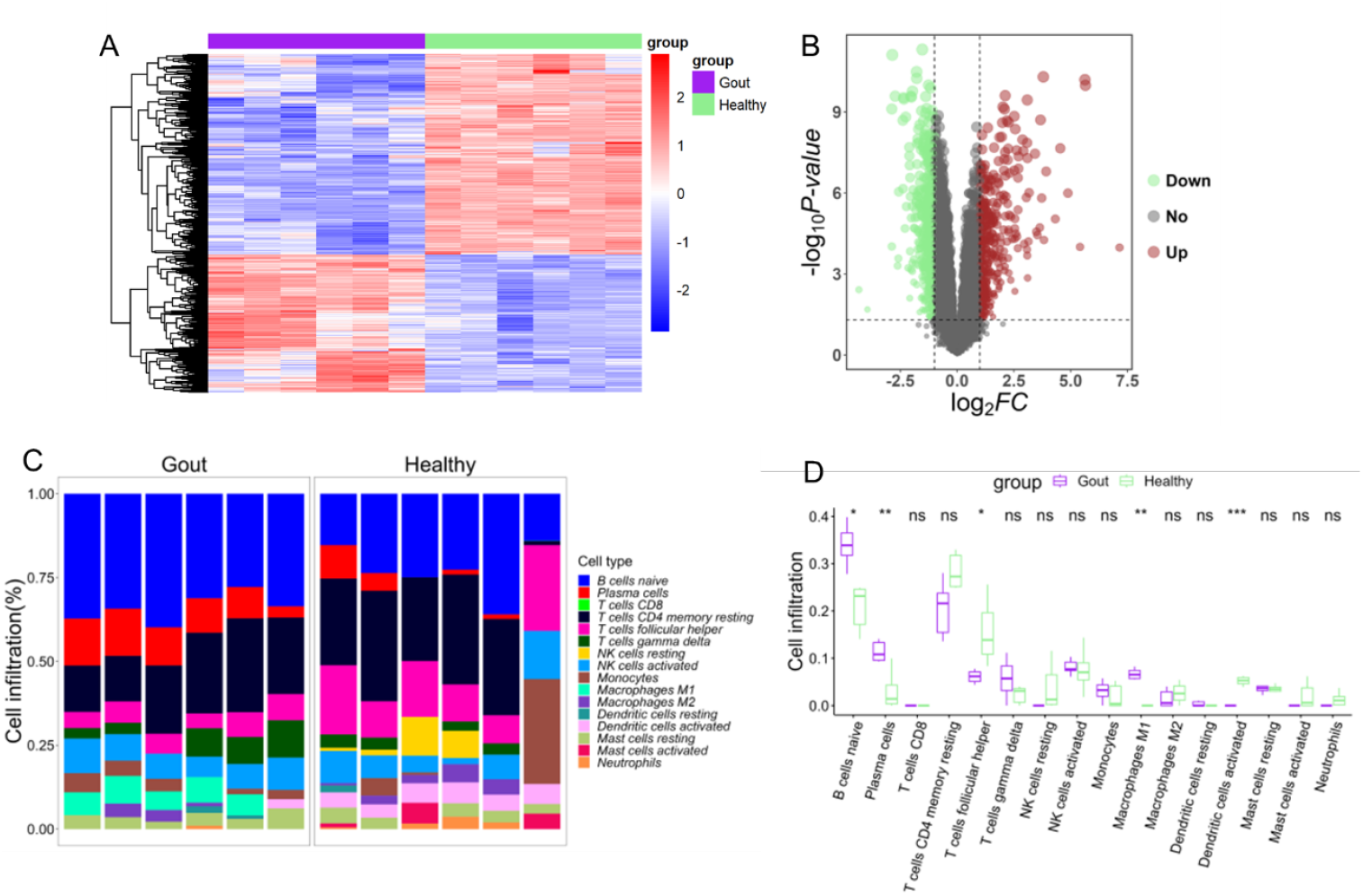
Results of bulk RNA-seq dataset analysis. A: Heatmap of DEGs between the disease and control groups in the GSE160170 dataset; B: Volcano plot of DEGs between the disease and control groups in the GSE160170 dataset; C: Bar chart showing immune cell infiltration across samples in the GSE160170 dataset; D: Box-and-whisker plot illustrating differences in immune cell infiltration between the disease and control groups in the GSE160170 dataset.

### 3.4 Identification of communication genes and pathway enrichment analysis

Intersection of macrophage marker genes from the single-cell transcriptomics dataset GSE211783 with DEGs from the bulk RNA-seq dataset GSE160170 yielded 102 intersecting genes (Figure 4A). GSVA pathway analysis on these 102 DEGs revealed14 differentially expressed signalling pathways, including the Wnt, TGF-β, JAK-STAT, Chemokine, mTOR, Calcium, and MAPK signaling pathways, as well as Ubiquitin-mediated proteolysis, Apoptosis, Cysteine and methionine metabolism, Cell cycle, Endocytosis, Lysosome and Glycolysis and gluconeogenesis. All 14 differentially expressed signaling pathways exhibited higher activity scores in the gout group (*P* < 0.05, Figure 4B). Figure 4C illustrates the PPI network of the 102 DEGs, in which the darker line colour denotes the higher interaction score. Core subnetworks were identified using the MCC, MNC, and DMNC algorithms, as shown in Figure 4D. The three core subnetworks collectively contained 25 core genes.

**Figure 4:**
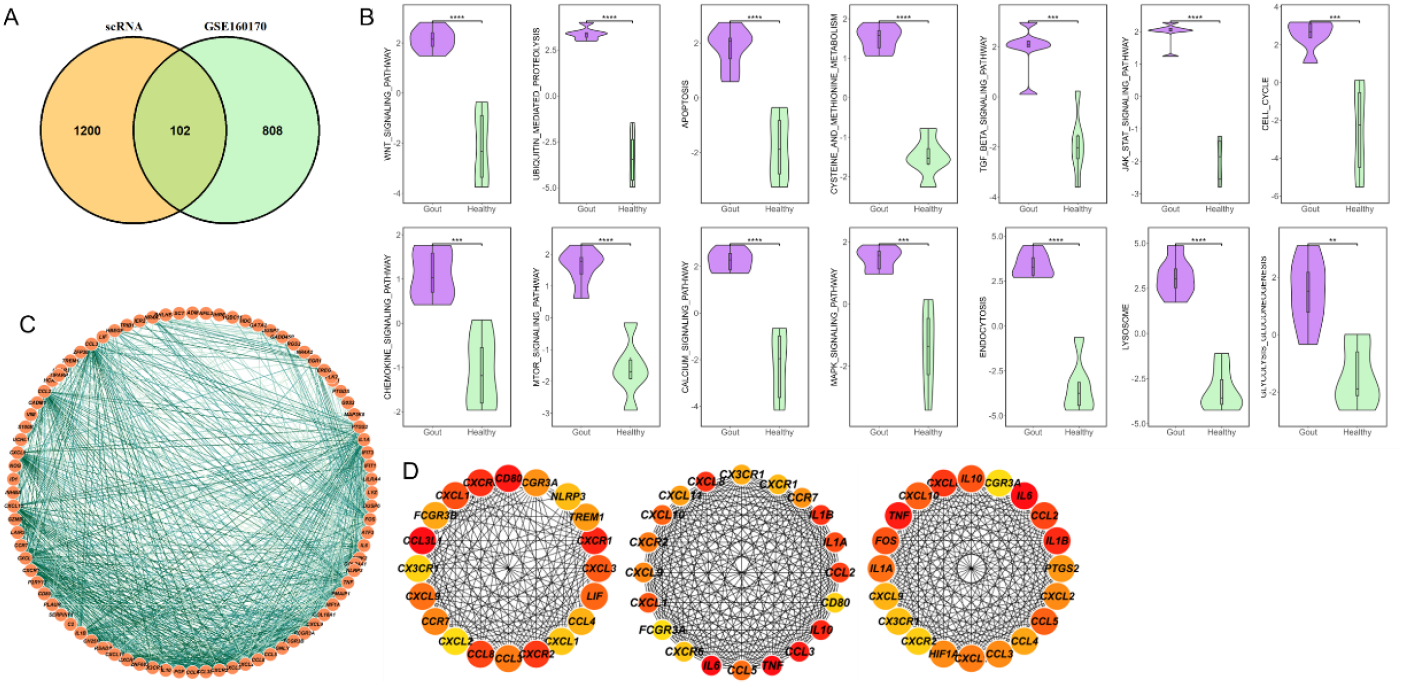
Identification of communication genes and pathway enrichment analysis. A: Venn diagram illustrating the intersection between DEGs from GSE160170 and macrophage marker genes from GSE211783; B: Activity scores of the 14 pathways identified by GSEA pathway analysis in disease and normal samples; C: The PPI network of 102 DEGs; D: PPI core subnetworks (calculated using the DMNC, MCC, and MNC algorithms, from left to right).

### 3.5 Differential expression results for core genes

Twenty-five genes showed significant differential expression between the disease and control groups (*P* < 0.05), as shown in Figure 5. Among these, TREM1, TNF, PTGS2, NLRP3, LIF, IL10, IL1B, IL1A, HIF1A, FOS, CXCL8, CXCL3, CXCL2, CXCL1, CD80, CCL3L1, and CCL3 exhibited markedly higher expression, whereas FCGR3B, FCGR3A, CXCR6, CXCR2, CXCR1, CX3CR1, CCR7, and CCL8 showed significantly lower expression in the gout group relative to the healthy group.

**Figure 5:**
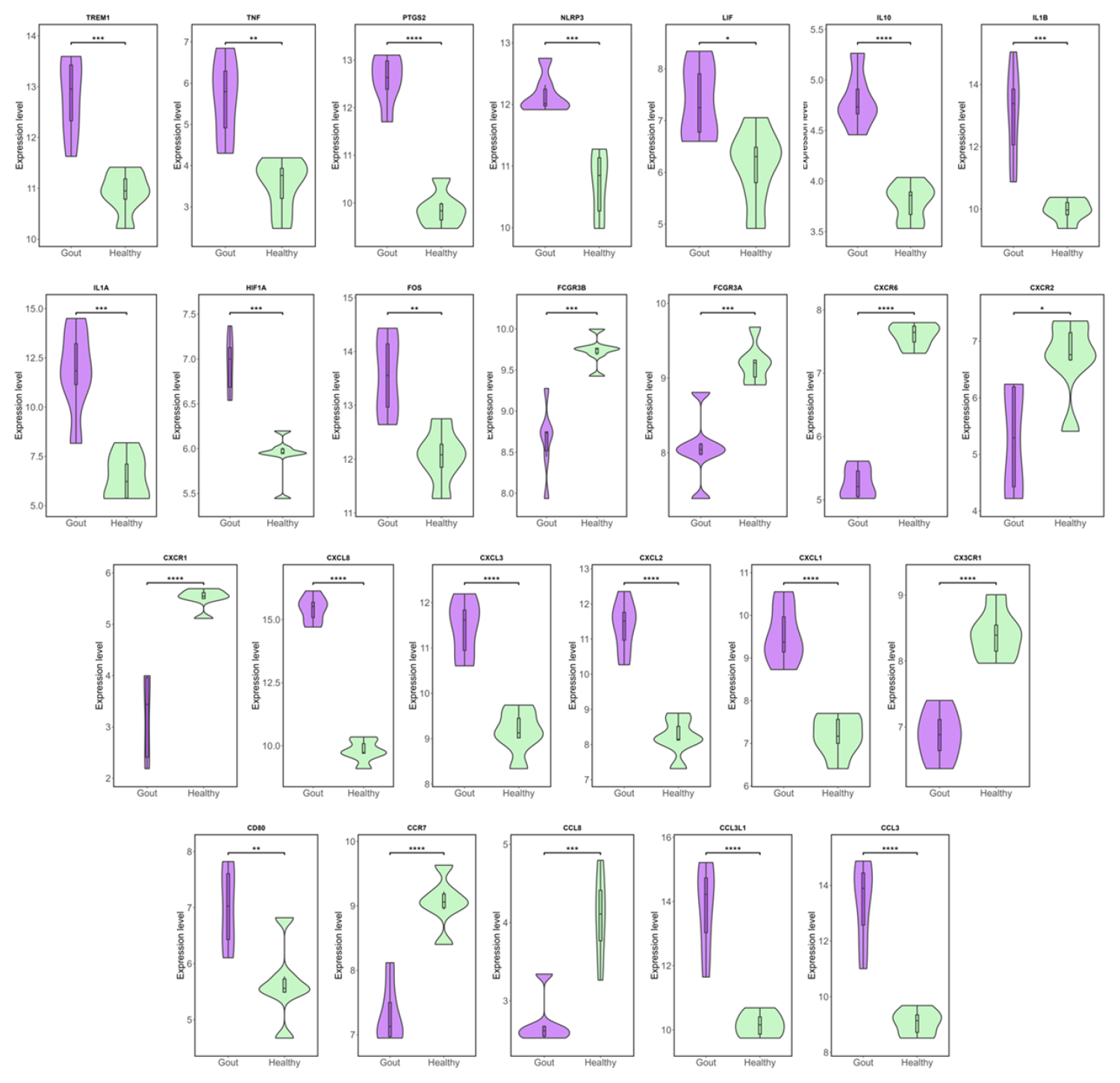
Violin plot of differential expression for 25 core genes (purple represents the gout group, green represents the healthy group)

### 3.6 Results of correlation analysis between core genes and macrophage infiltration

All 25 core genes were significantly associated with macrophage infiltration (P < 0.05); specific results are shown in Figure 6. Among these, TREM1, TNF, PTGS2, NLRP3, LIF, IL10, IL1B, IL1A, HIF1A, FOS, CXCL8, CXCL3, CXCL2, CXCL1, CD80, CCL3L1, and CCL3 were positively correlated with macrophage infiltration, whilst FCGR3B, FCGR3A, CXCR6, CXCR2, CXCR1, CX3CR1, CCR7, and CCL8 were negatively correlated with macrophage infiltration.

**Figure 6.**
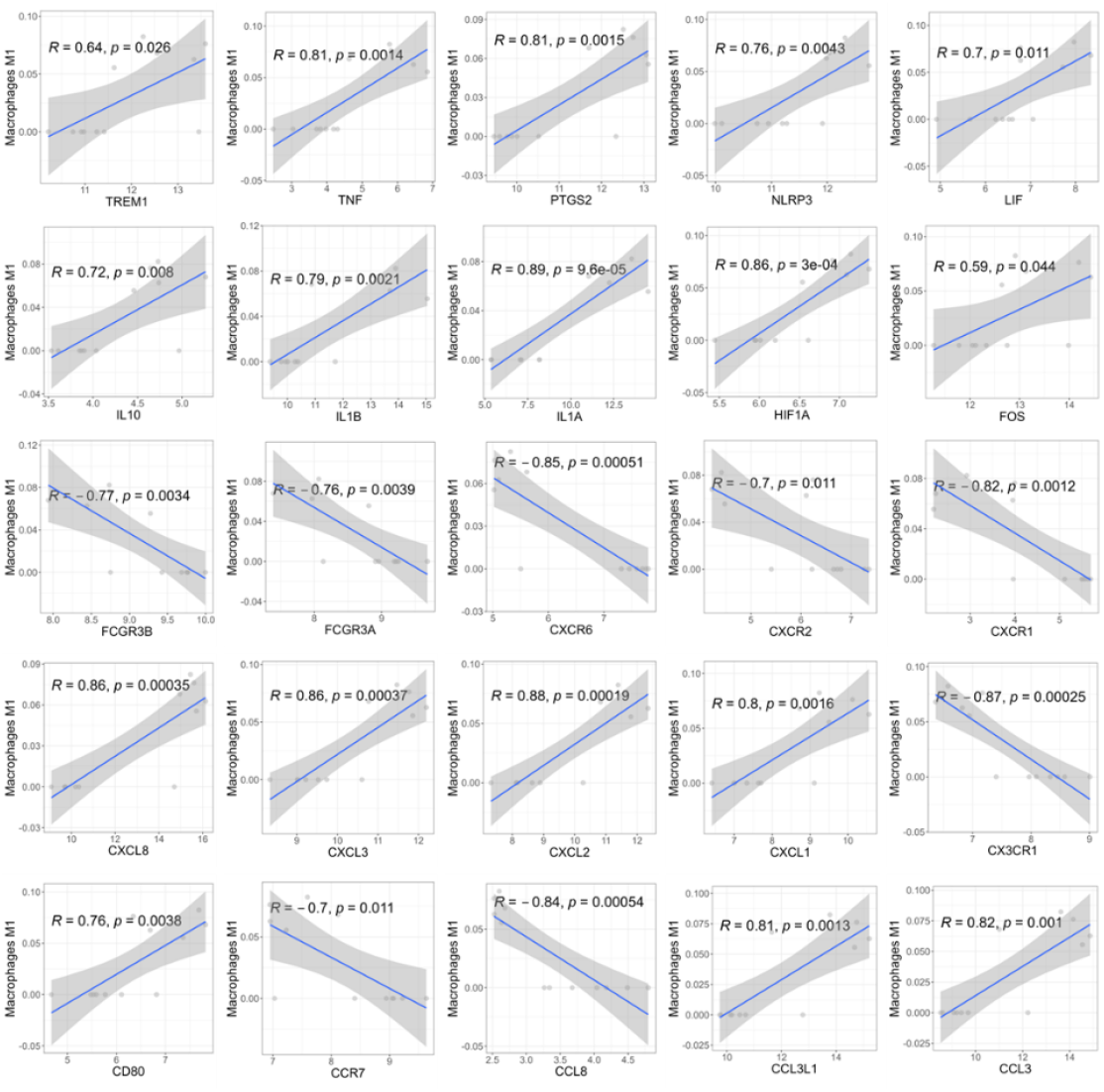
Correlation analysis between 25 core genes and the level of macrophage infiltration

### 3.7 Results of the correlation analysis between core genes and drug sensitivity

Figure 7 illustrates the correlation between the 25 core genes and drug sensitivity based on correlation analysis. Positive correlations are shown in red, negative correlations in blue, and dot size reflects the FDR value, with larger dots indicating lower FDR. Among these, TREM1, CCL8, and CXCR6 showed a weaker correlation with drug sensitivity.

**Figure 7.**
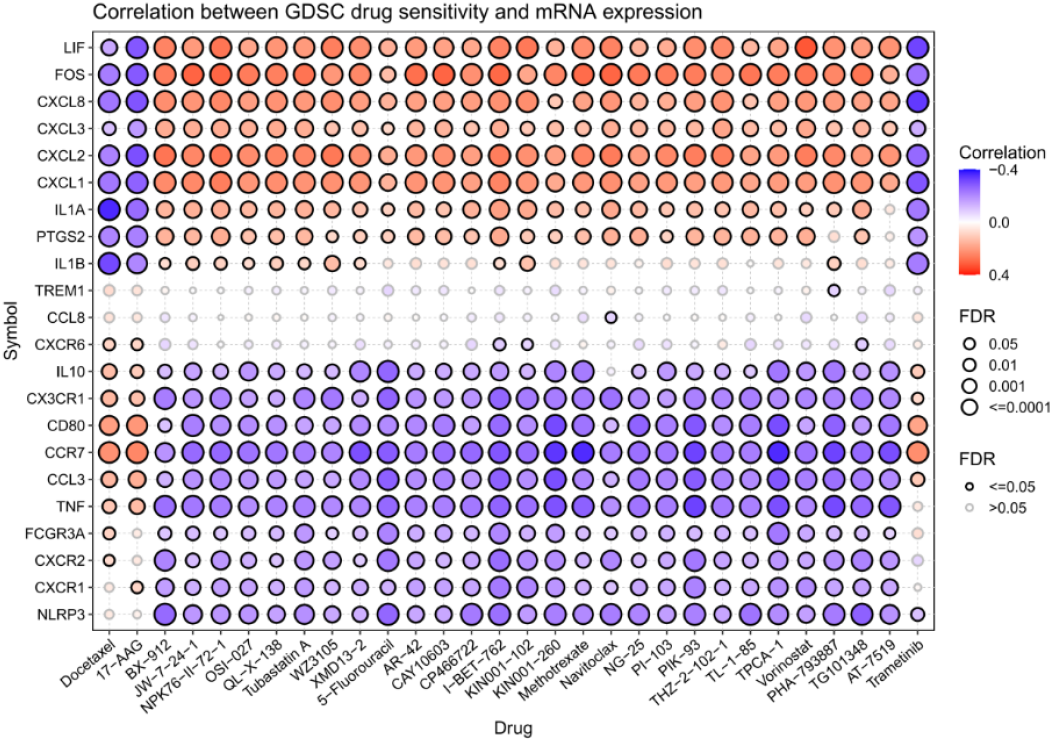
Correlation analysis between core genes and drug sensitivity (IC_50_)

### 3.8 Correlation analysis between TREM1 and signal pathway scores

TREM1 was selected from the 25 core genes for further investigation. The results of the analysis of TREM1 against 14 signaling pathway scores are shown in Figure 8. TREM1 showed strong correlations with the Wnt, TGF-β, JNK/STAT, chemokine, mTOR, calcium ion, and MAPK signaling pathways, as well as ubiquitination-mediated pathway, apoptosis pathway, glycolysis pathway, cell cycle pathway, endocytosis pathway, lysosomal pathway, and threonine and methionine-mediated pathways.

**Figure 8.**
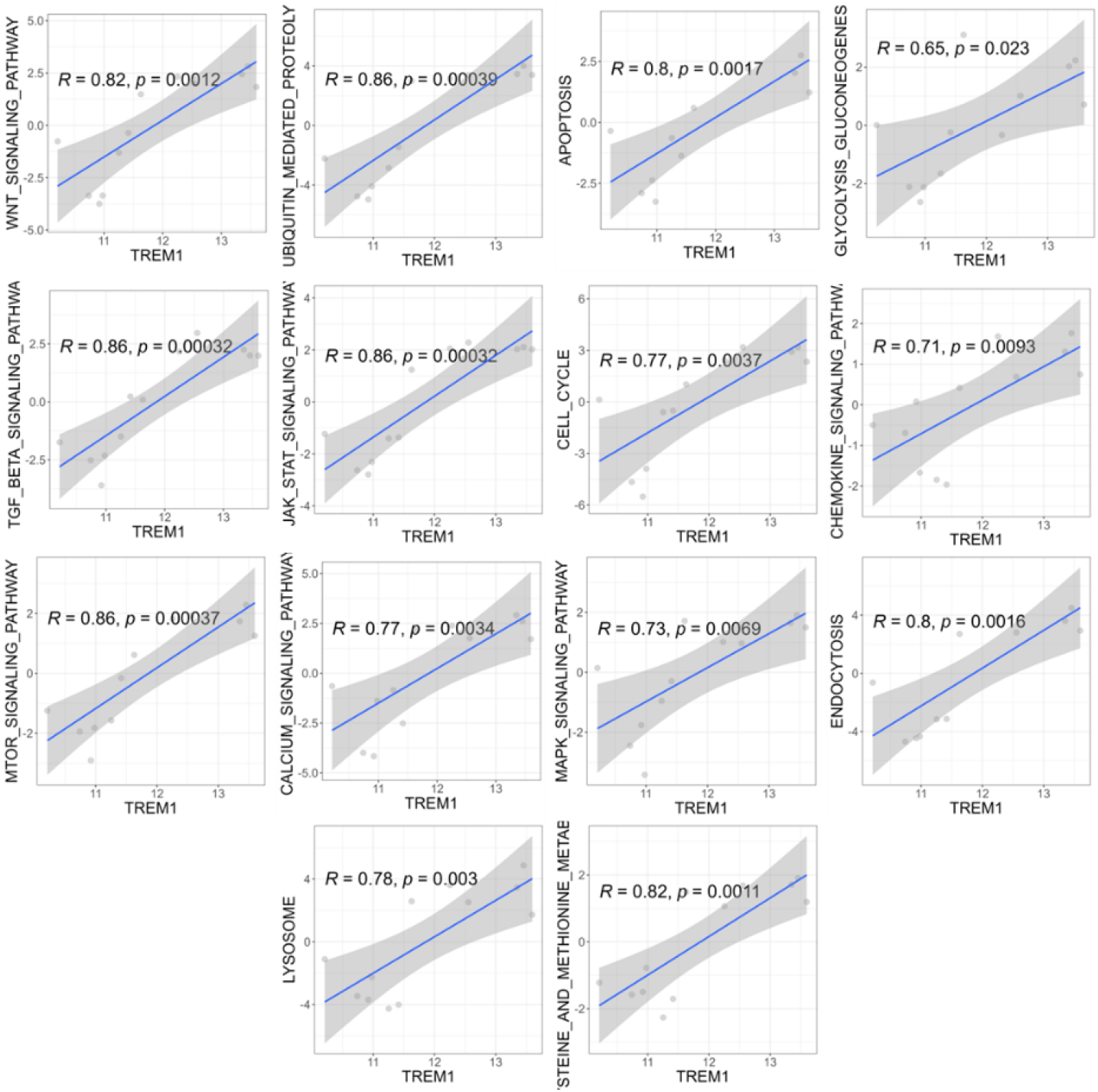
Correlation analysis between TREM1 and pathway scores

### 3.9 Expression of TREM1 in the joint tissues of mice with GA

The right hind paw pad swelling index in the Model group was significantly higher than that in the Control group (Figure 9A). HE staining revealed that, compared with the Control group, the joint tissues of mice in the Model group exhibited marked synovial oedema, synovial cell proliferation, inflammatory cell infiltration, and neovascularisation (Figure 9B). IHC results showed an increase in TREM1-positive cells in mouse joint tissues from the Model group (Figure 9C). WB analysis revealed markedly elevated TREM1 protein expression in mouse joint tissues from the Model group (Figure 9D).

**Figure 9.**
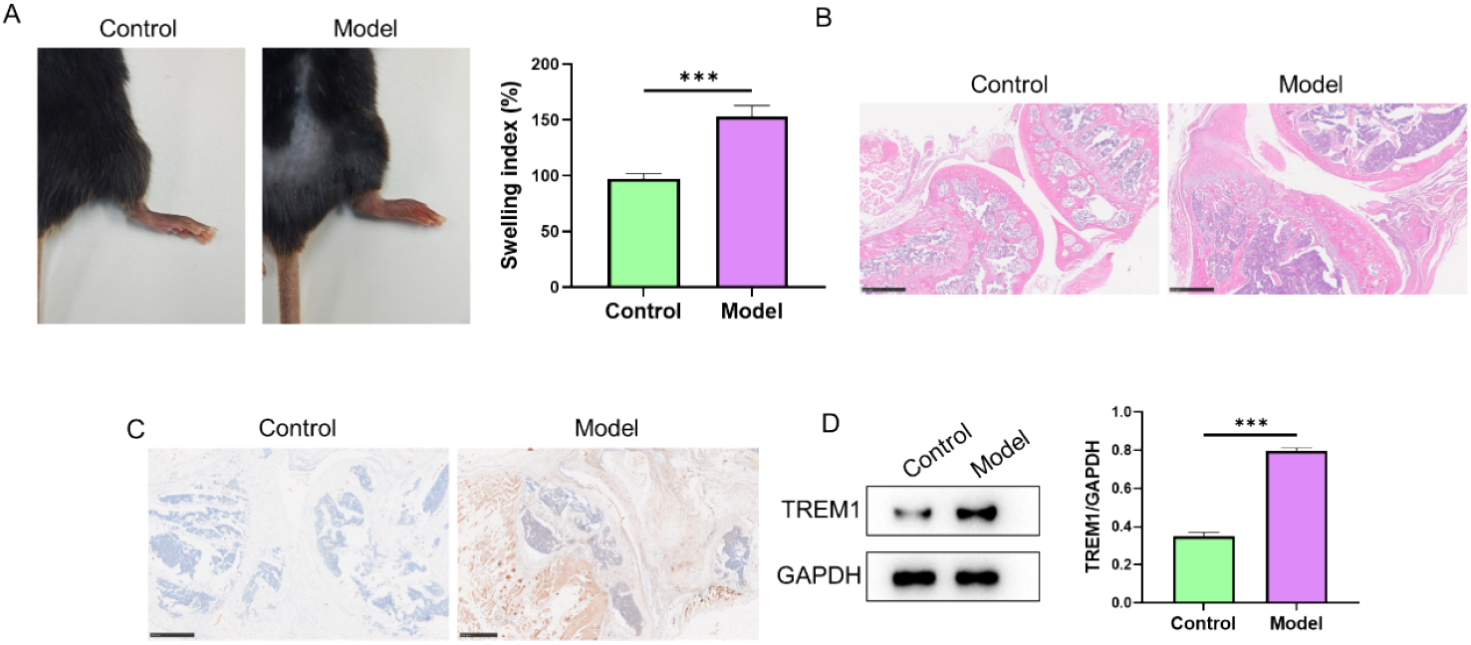
Expression of TREM1 in the joint tissues of mice with GA. A: Joint swelling index in mice from each group; B: Morphology of mouse joint tissue observed via HE staining; C: IHC-based evaluation of TREM1 expression in mouse joint tissue; D: WB-based quantification of TREM1 expression in mouse joint tissue. ***P < 0.001

## 4. Discussion

GA is a common metabolic arthritis marked by the accumulation of uric acid crystals in both joint and non-joint tissues, which triggers acute arthritis. Clinically, patients with GA commonly report severe nighttime joint pain, accompanied by redness, swelling, and a burning sensation in the affected joints. Progression of the disease may lead to the involvement of additional joints, including the knees and ankles ^[18]^. A deeper exploration of the specific molecular mechanisms involved in GA initiation and progression is crucial for identifying effective treatments and essential for the management and prevention of GA. Over the past few decades, with advances in technology, an increasing number of studies have begun to employ bioinformatics methods to conduct in-depth research into GA. Bioinformatics encompasses fields such as genomics, gene regulation, proteomics, biological networks and metabolomics, utilizing computer technology to analyze and process vast amounts of biological data, thereby helping researchers to better understand disease mechanisms^[19]^ .

To date, transcriptomic and single-cell sequencing approaches have been widely used to identify candidate biomarkers implicated in gout progression. Xinyi Wang et al. performed weighted gene co-expression network analysis to screen for candidate biomarkers for gout. Their results indicated that IL-1β, IL-6, CXCL1, CXCL2, and CXCL8 serve as key hub genes within the PPI network. Marked differences in CXCL1, CXCL2, and CXCL8 expression were identified between the gout and healthy control groups, and 10 chemical compounds were predicted to be associated with these proteins ^[20]^. Proteomic analysis by Chengjin Lu et al. revealed 40 differentially expressed proteins, which showed predominant enrichment in the following pathways: the complement and coagulation cascades, lysosomal function, autophagy, and purine metabolism. They provided evidence from in vivo and in vitro studies that exosomes are involved in GA initiation and progression through regulation of lysosomal dysfunction and purine metabolism ^[21]^. By integrating bioinformatics and machine learning results, Hui-Li Han et al. characterized B3GALT2 as a key gene exhibiting downregulated expression in GA. Furthermore, experimental findings from in vitro and in vivo models suggested that the TFAP2A/B3GALT2 axis is crucial for protecting against GA via suppression of pyroptosis mediated by the NLRP3 inflammasome ^[22]^.Wei Wang et al. identified nine metabolites, distinct from those associated with other inflammatory conditions, through metabolomics combined with GC/LC-MS analysis. These metabolites show potential as biomarkers for the diagnosis and treatment of GA; notably, kynurenic acid, thymine and phosphatidylinositol were found to be contrary to trends observed in previous studies ^[23]^. Isidoro Cobo et al. demonstrated, through whole-genome transcriptomic and biochemical analyses, that the metabolic inflammatory transcriptional programmes induced by MSU in human and mouse macrophages are significantly distinct from those triggered by LPS. MSU specifically induces the upregulation of genes associated with lipid and amino acid metabolism, glycolysis, and SLC transporters, which results in metabolic remodeling in the serum of GA patients and mice. From a mechanistic perspective, the inflammatory metabolic changes observed in GA may be regulated through sustained JUN expression and its enhanced binding to target gene promoters via JNK signaling ^[24]^ .

It is evident, therefore, that the specific molecular regulatory pathways involved in the onset and progression of GA are highly complex and remain poorly understood. The application of transcriptomic and cellular analysis techniques to whole synovial tissue has identified specific cell populations associated with various forms of GA ^[25]^. However, most studies have focused on pre-selected cell types, examined whole tissues rather than cell fractions, or utilized a single technical platform. Recent advances in single-cell technologies enable high-resolution and objective identification of disease-related cell subpopulations in human tissues ^[26]^. In the current research, through screening of both the single-cell dataset GSE211783 and the bulk RNA-seq dataset GSE160170, we identified 102 DEGs. GSVA-based pathway analysis of these identified DEGs revealed 14 differentially expressed signaling pathways: including the Wnt, TGF-β, JAK-STAT, Chemokine, mTOR, Calcium, and MAPK signaling pathways, as well as Ubiquitin-mediated proteolysis, Apoptosis, Cysteine and methionine metabolism, Cell cycle, Endocytosis, Lysosome, Glycolysis,and gluconeogenesis. Importantly, all these pathways exhibited higher activity scores in the gout patient group. A PPI network derived from the 102 DEGs was analyzed, and core subnetworks were identified using the MCC, MNC and DMNC algorithms; these three core subnetworks collectively contained 25 core genes. Expression levels of these 25 genes differed markedly between the disease and control groups. Among them, TREM1, TNF, PTGS2, NLRP3, LIF, IL10, IL1B, IL1A, HIF1A, FOS, CXCL8, CXCL3, CXCL2, CXCL1, CD80, CCL3L1, and CCL3 exhibited markedly higher expression, whereas FCGR3B, FCGR3A, CXCR6, CXCR2, CXCR1, CX3CR1, CCR7, and CCL8 demonstrated significantly lower expression in the gout group relative to the healthy group. A significant correlation was found between macrophage infiltration levels and the expression of all 25 genes. Among these 25 genes, TREM1, CCL8, and CXCR6 showed a weaker correlation with drug sensitivity. TREM1 was selected as the target gene for this study for subsequent investigation. TREM1 exhibited a strong correlation with 14 differentially expressed signaling pathways.

Among the 25 differentially expressed core genes identified in this study, CXCL8 is a classic neutrophil chemokine; CXCL3, CXCL2, and CXCL1 also belong to the CXC family and primarily chemotactically attract neutrophils ^[27]^. CCL3L1, CCL3, and CCL8 are cytokines belonging to the CC family, primarily involved in the chemotaxis of macrophages, T cells, monocytes and other cells ^[28]^. IL1B and IL1A are classic pro-inflammatory factors that mediate systemic inflammatory responses ^[29]^.TNF is also a classic pro-inflammatory factor that plays a key role in diseases such as rheumatoid arthritis ^[30]^. LIF is a multifunctional cytokine involved in the regulation of inflammatory homeostasis ^[31]^. IL-10 is a classic anti-inflammatory factor involved in immune suppression in inflammation-related diseases ^[32]^. NLRP3 is the core sensor of the inflammasome, responsible for the activation and modulation of downstream inflammatory factors, including IL-1β ^[33]^. PTGS2, HIF1A, and FOS are key proteins mediating inflammatory responses via different pathways, regulating the expression of various genes or proteins associated with inflammatory responses ^[34-36]^. CD80 is a key ligand for T-cell activation during the inflammatory response ^[37]^. CXCR6, CXCR2, CXCR1, CX3CR1, and CCR7 are all chemokine receptors that act in concert to mediate the recruitment, retention and lymph node homing of neutrophils, plasma cells, monocytes, T cells, dendritic cells, and natural killer cells ^[38]^. FCGR3B and FCGR3A encode receptors on immune cell surfaces, mediating antibody-dependent cell-mediated cytotoxicity and phagocytosis, and participating in inflammatory responses ^[39, 40]^. Ultimately, TREM1, which is expressed in macrophages, was ultimately chosen as the target gene for further analysis in this study.

TREM1 has a well-established role in inducing macrophages and neutrophils in various inflammation-related diseases; however, its involvement in GA remains insufficiently characterized. C. E. Collins et al. reported elevated TREM1 expression in patients with septic arthritis or rheumatoid arthritis ^[41]^. Jennifer Lee et al. examined 37 samples from patients with gout and showed, through FACS analysis, real-time RT-PCR, and immunostaining, that MSU-induced elevated TREM1 expression correlates with inflammation in acute GA ^[42]^. Yonglong He et al. reported that TREM1 may aggravate MSU-triggered acute inflammation, and that inhibition of TREM-1 may represent a novel therapeutic strategy for alleviating acute GA ^[43]^.Yair Molad et al. found that inhibiting TREM-1 alleviates GA by selectively suppressing pro-inflammatory cytokines and chemokines, without affecting TGF-β production ^[44]^. Chao-Nan Zhang et al. demonstrated that electroacupuncture treats GA by downregulating TREM-1 expression in synovial tissue ^[45]^. This study validated abnormally high TREM1expression in mouse joint tissues within a mouse model of GA, which aligns with the findings of prior reports.

In summary, analysis of the single-cell dataset GSE211783 and the bulk RNA-seq dataset GSE160170 in this study revealed 102 DEGs between GA and normal samples. These DEGs were enriched in 14 differentially expressed pathways. Following the construction of a PPI network for these 102 DEGs, 25 key DEGs were identified within a key subnetwork. Among these, TREM1, TNF, PTGS2, NLRP3, LIF, IL10, IL1B, IL1A, HIF1A, FOS, CXCL8, CXCL3, CXCL2, CXCL1, CD80, CCL3L1, and CCL3 exhibited markedly higher expression, whereas FCGR3B, FCGR3A, CXCR6, CXCR2, CXCR1, CX3CR1, CCR7, and CCL8 showed significantly lower expression in the gout group relative to the healthy control group. Significant associations were observed between macrophage infiltration levels and all 25 DEGs. TREM1 was chosen as the target gene for further analysis in this study. TREM1 showed strong correlations with 14 differentially expressed signaling pathways. Furthermore, this study detected higher levels of abnormal TREM1 expression in GA mouse samples, consistent with the results of bioinformatics analysis. Therefore, TREM1 may represent a biological target for both the clinical diagnosis and treatment of GA. Several limitations of this study need to be taken into consideration. Firstly, the expression of TREM1 was validated only at the in vivo level in mice; further clinical samples need to be collected to verify TREM1 expression at the clinical level. Secondly, this study has merely validated the expression of TREM1; additional studies are needed to clarify the specific molecular mechanisms responsible for the alleviative effects of TREM1 on GA, and to validate these findings at the animal, cellular, molecular, and even clinical levels, thereby providing more robust evidence to support the feasibility of TREM1 as a therapeutic and diagnostic biological target in GA in clinical settings.

## Data availability

Publicly available datasets were analyzed in this study. This data can be found here: [Gene Expression Omnibus, https://www.ncbi.nlm.nih.gov/geo/]

## Author contributions

Wei Jinfeng drafted the manuscript,Qiu Hongbin revised the manuscript. Wei Jinfeng and Zheng Jiarui drew Figs. 1 to 9. All authors reviewed and approved the final manuscript.

## Funding

Project of the Natural Science Foundation of Heilongjiang Province (NO.PL2025H045). Basic Research Expenses of Provincial Colleges and Universities of Heilongjiang Province (NO.2025-KYYWF-ZR0673).

Basic Research Expenses of Provincial Colleges and Universities of Heilongjiang Province (NO.2023-KYYWF-0608).

## Declarations

### Consent for publication

Not applicable.

